# COSSMaT Study: Protocol for Developing a Core Outcome Set for Severe Malaria Treatment Trials

**DOI:** 10.1101/2025.05.05.25326984

**Authors:** Gideon D Asamoah, Sharon Love, Diana M Gibb, Kathryn Maitland, Elizabeth C George

## Abstract

**Objectives:** Severe malaria remains a significant global health issue, especially in endemic regions, due to its high rates of morbidity and mortality. Despite progress, the variability in outcome measures across clinical trials hampers the synthesis of evidence and the development of standardised treatment guidelines. Developing a Core Outcome Set (COS) for severe malaria trials is crucial to address these inconsistencies, enhance research quality, and support evidence-based clinical practices. This study aims to develop a core outcome set for trials in the treatment of severe malaria.

**Method:** The study will follow the COMET Initiative guidelines to develop a reliable, valid, and responsive COS for clinical trials focused on severe malaria treatment. This will involve updating a systematic review of outcomes reported in severe malaria trials and collecting outcomes reported by patients, parents, and guardians/caregivers through qualitative research. Subsequently, collaboration with key stakeholders with expertise in severe malaria (including patient/support group representatives, healthcare workers, pharmaceutical industry representatives, policymakers, and researchers) will prioritise outcomes through a multi-stage Delphi survey. A Consensus meeting will be held virtually with stakeholder representatives to discuss and vote on the outcomes to include in the final COS.

**Conclusion:** The global burden of malaria, particularly among children and vulnerable populations such as pregnant women, underscores the necessity of a COS to standardise outcomes in severe malaria trials. Addressing this gap is essential for standardising definitions, enhancing research quality, and facilitating transparent and credible comparisons. Establishing a COS not only improves individual studies and the combination of studies but also contributes to evidence-based treatment protocols to ensure effective treatment is identified, ultimately reducing the global health burden of severe malaria.

**Study registration:** Core Outcome Measures in Effectiveness Trials (COMET) initiative 2908. https://www.comet-initiative.org/Studies/Details/2908

## Introduction

Malaria persists as a formidable global health challenge, exacting a heavy toll on both morbidity and mortality rates worldwide (1–3). Africa, in particular, bears a disproportionate burden, with 94% of malaria cases (246 million) and 95% (569,000) of malaria deaths recorded by the World Health Organization (WHO) in 2023 (2). Children below the age of five constituted approximately 76% of malaria-related fatalities, making it a leading cause of childhood illness and death in the region (2,4). Beyond its impact on children, malaria significantly influences directly maternal and indirectly neonatal outcomes, since it heightens the risks of abortion, low birth weight, preterm delivery, stillbirth, and neonatal death (5–7).

The pursuit of improved malaria treatment remains an ongoing endeavour, particularly for severe disease(8,9). However, a significant impediment arises from the lack of consensus on appropriate outcomes for use in clinical trials, posing a challenge in measuring and evaluating the efficacy of antimalarial and/or adjunctive treatments. A systematic review has highlighted the considerable heterogeneity, with 101 diverse outcome measures reported in 27 trials spanning from 2010 to 2020 (10). This disparity has led to the measurement of diverse outcomes and the utilisation of various instruments for assessing the same outcome, resulting in inconsistencies in reported results and complicating the comparison of outcomes in systematic reviews and meta-analyses (10). Creating a Core Outcome Set (COS) for severe malaria treatment in trials could help mitigate the current deficiencies in addressing reporting challenges.

COS is defined by COMET as “an agreed standardised set of outcomes that should be measured and reported, as a minimum, in all clinical trials in specific areas of health or health care.”(11) While a previous study has developed COS for malaria in the context of vaccine trials (12), a notable gap exists concerning the treatment of severe malaria, especially with the inclusion of adjunctive therapies (10). This void is a critical concern for global health, given the severe and widespread impact of severe malaria. Urgent attention is warranted to address this gap.

The scope of the COS specifies the particular healthcare domain or condition it addresses, detailing its application’s target population, interventions, and settings. This clarity ensures its appropriate and consistent use, thereby avoiding any ambiguity (13–15). For this COS, the scope encompasses severe malaria, characterised by complications such as impaired consciousness, acidosis, and severe anaemia. The target population includes all groups affected by severe malaria, including children under five, adults, pregnant women, and travellers who reside in endemic and non-endemic regions. The interventions involve antimalarial treatments and adjunctive therapies to manage complications. The setting is within clinical trials to standardise outcome selection and reporting, ensuring relevance and consistency across studies.

## Methods

This study protocol will adhere to the guidelines outlined in the Core Outcome Set- STAndardised Protocol Items (COS-STAP) Statement checklist (22). The study has been registered on the COMET Initiative website (https://www.comet-initiative.org/Studies/Details/2908). The study will follow the COMET Initiative guidelines for developing a core outcome set to produce a reliable, valid, and responsive minimum set of outcomes to be measured and reported in all clinical trials considering the treatment of severe malaria (13).

### Study summary

We conducted a search for an existing COS for severe malaria treatment trials, adhering to the COMET guideline’s recommendation to prevent duplication of ongoing or pre-existing studies. No such study was found.

We will develop the COS through the following phases:

- Phase 1: To develop a “long list” of outcomes to guide the creation of a core outcome set for severe malaria treatment trials, we will use the following information sources:
  1. Systematic review update: Identify additional outcome sets reported for trials in the treatment of severe malaria.
  2. Consultation with clinical experts in severe malaria to incorporate recent and unpublished research insights.
  3. Qualitative study: Identify severe malaria outcomes reported by patients, parents, and guardians/caregivers for measurement in treatment trials.
- Phase 2: Develop a preliminary COS (informed by the “long list” of outcomes from phase 1) with key stakeholders through a multi-stage (two or three round web-based) Delphi survey.
- Phase 3: Hold a consensus meeting with stakeholders to discuss and agree on the final COS for severe malaria treatment.
- Phase 4: Develop a dissemination and implementation strategy for the final COS.

### Phase 1: “Long list” of outcomes to guide the creation of a core outcome set for severe malaria treatment trials

#### Systematic review update

This will involve updating the previous systematic review on outcomes reported in trials for severe malaria treatment between 1 January 2010 and 30 July 2020 (10). This review included 27 trials out of 282 screened, involving 10,342 patients from 19 countries, with 70% from Africa and 30% from Asia, reporting a total of 101 distinct outcome measures. The update aims to include any newly published or registered trials in severe malaria from 31 July 2020 onwards. The systematic review will follow the guidelines outlined in the “Reporting Systematic Reviews and Meta-Analyses of Studies That Evaluate Health Care Interventions” checklist (16). These systematic processes, detailed in the previous study (10), includes establishing eligibility criteria, identifying information sources, formulating an electronic search strategy, specifying the study selection process, deciding on data collection procedures and elements, and describing summary measures and analysis methods.

#### Consultation with clinical experts

Clinical experts will be engaged to provide essential insights into practical and clinically relevant outcomes. Additionally, these consultations may help identify outcomes not captured in the systematic review.

#### Qualitative study

The COMET guidelines emphasise the importance of involving patients and the public throughout the development process to ensure that selected outcomes are relevant and meaningful to those directly impacted (13). Such engagement ensures that all potentially relevant outcomes are considered, enhancing the comprehensiveness and relevance of the outcome list (17,18).

While other studies have reviewed qualitative studies on outcomes reported by patients, parents, and guardians (19,20), no study has focused on outcomes reported by these groups in severe malaria. We will employ a qualitative research methodology to explore the narratives shared by patients, parents/guardians, and caregivers about their experiences with severe malaria. This approach aims to capture their experiences and identify the outcomes they consider crucial for assessment in severe malaria treatment trials. We plan to recruit participants from Low- and Middle-Income Countries (LMICs), particularly in Sub-Saharan Africa, where the burden of malaria is disproportionately high (21–23). This strategic recruitment aims to ensure representation from regions facing significant challenges associated with severe malaria, providing a comprehensive and contextually relevant understanding of the impact and experiences related to this condition. The qualitative study will adhere to the Consolidated Criteria for Reporting Qualitative Research (COREQ) guidelines (24) and the reporting recommendations for qualitative research methods in COS development outlined by Jones et al. (17,25).

#### Generation of the “long list” of outcomes

The long list will encompass all potentially relevant outcomes from diverse information sources, including systematic literature reviews, clinical experts and interviews with patients, parents, and guardians (26–29). Once these outcomes are identified, similar measures across the various sources will be consolidated and systematically categorised into coherent domains. This process will utilise the outcomes framework for medical research recommended by the COMET initiative (30), other studies (10,31) and consultation with clinical experts.

### Phase 2: Delphi survey with stakeholders

The purpose of a Delphi survey is to collect opinions from a panel of participants including experts in severe malaria and clinicians who regularly manage patients with severe malaria on a specific topic to achieve group consensus (32). Key stakeholders will be presented with plain language explanations of the “long list” of outcomes, and will then rate the importance of each item for inclusion in the COS through a multi-stage Delphi survey.

#### Stakeholder Groups and Recruitment

Key stakeholders, including patient/support group representatives, healthcare workers, pharmaceutical industry representatives, policymakers, and researchers, will be involved in the COS development process. Stakeholders will be identified and recruited through expert networks associated with public health agencies, regulatory authorities, research institutions, pharmaceutical industries, and civil society organisations. Notable examples of such networks include WHO, Malaria Consortium, Severe Malaria Africa - A Consortium for Research and Trials (SMAART) Consortium, the COMET Initiative, Novartis, Cipla, support groups representing individuals with lived experiences, and first authors of related published articles. We aim to promote the study and recruit stakeholders by developing a project webpage, featuring in network newsletters, presenting at seminars and conferences, using social media, and distributing leaflets. Participant information leaflets (PIS) and consent forms will be provided electronically.

#### Multi-stage Delphi Survey

This survey will consist of two (2) rounds using the long list of outcomes from the information sources (13), each separated by a 4-week interval. Participants will have 4 weeks to complete each survey, with the option to save and resume their responses later. A third round may be considered depending on the analysis of the first two rounds. Descriptive statistics will be used to analyse overall scores for each stakeholder group, ultimately determining inclusion or exclusion of outcomes.

In round 1, participants will rate the importance of outcomes for inclusion in the COS using a 9- point scale, categorised as 9-7 (critically important), 6-4 (important but not critical), and 3-1 (not important). They can also briefly justify their choices, suggest changes, and propose additional items not covered in the survey. Suggested outcomes from the first round will be reviewed by the study committee for inclusion in the second round. Only responses from participants who rated at least 50% of items will be included.

In round 2, a summary of overall scores and scores from stakeholder groups on each outcome will be provided to participants along with their own scoring. Participants will be aware of others’ views but won’t know individual scores to maintain anonymity, allowing unbiased expression of opinions. Participants can maintain or adjust their ratings in the second round if desired. Each round will have a 4-week completion timeframe.

#### Consensus definition

Outcomes that receive scores of 7–9 from 70% or more participants and scores of 1–3 from less than 15% of participants across all stakeholder groups will be considered for inclusion in the COS during the consensus meetings. Outcomes will be excluded if they are scored between 1 and 3 by at least 70% of participants across all stakeholder group and between 7 and 9 by less than 15%. Outcomes that do not fall into either category will be classified as having no consensus and discussed at the consensus meeting, as applied in other COS studies (20).

### Phase 3: Consensus meetings

All Delphi participants will be invited to join an online consensus meeting via Zoom. Each stakeholder group will be represented by at least two participants. If an invited participant cannot attend, efforts will be made to find a suitable replacement from the same stakeholder group. To facilitate broad international participation, meetings will be scheduled to accommodate different time zones and participant availability.

Prior to the meetings, all necessary materials (outcomes rated through the Delphi process and voting patterns from different stakeholder groups) will be circulated to ensure participants are well-prepared. To enhance engagement and decision-making, participants will be provided with a secure weblink to cast their votes remotely during the meeting.

The primary objective of the consensus meeting is to finalise the Core Outcome Set (COS) through structured discussions involving a diverse international panel. Outcomes identified through the Delphi process will be reviewed alongside voting patterns from different stakeholder groups. Discussions will be chaired by experienced, non-voting facilitators to ensure neutrality. Following these discussions, participants will engage in anonymous, computerised voting. An outcome will be included in the final COS if it receives at least 70% approval from participants, with representation from all stakeholder groups.

### Phase 4: Dissemination strategy

The result of this research will be written as part a PhD dissertation, publication(s) and disseminated at conferences, on the project webpage, featured in network newsletters, presented at seminars, as well as distribution of leaflets to all expert networks associated with the treatment of severe malaria in trials. To enhance accessibility, a plain English summary will be provided in infographics and on social media to facilitate broader engagement. The study protocol and results will be publicly available on Open Science Framework to facilitate free and widespread sharing of the research. Participants will receive the findings of this research if they consent to it.

## Discussion and Conclusion

To the best of our knowledge, there is currently no established COS for trials specifically focused on severe malaria treatment. We aim to develop a COS for severe malaria trials to standardise the outcomes reported in such trials. A COS for severe malaria treatment would provide a framework, ensuring precise and consistent definitions of outcomes. This standardisation is crucial for facilitating comparisons across diverse studies and improving the overall quality of research in this critical area. By addressing this challenge, researchers can enhance the transparency of their findings, reduce outcome reporting bias and research waste, and strengthen the credibility and robustness of future trials (26,33,34).

Moreover, establishing a COS for severe malaria not only contributes to the refinement of individual studies but also aids in the selection of a single valid and reliable outcome measurement instrument for each outcome, facilitating more comprehensive meta-analyses within the broader malaria research domain (10,34,35). This, in turn, supports the development of evidence-based treatment protocols, providing a solid foundation for clinical practices. In essence, addressing the current gap by developing a COS for severe malaria treatment is key to advancing research, improving patient outcomes, and helping to reduce the global health burden posed by severe malaria.

### Study status

Protocol version 1.0, April 2025.

The systematic review for phase one has been completed, while the qualitative study is still in progress. Recruitment for phase two participants has not started yet, but it is anticipated to be finished by August 2025.

## Data Availability

All data produced in the present study are available upon reasonable request to the authors.

## Availability of data and materials

The final core outcome set will be submitted for peer review and reported according to the Core Outcome Set-Standards for Reporting (COS-STAR).

## Author Contributions

GDA serves as the chief investigator, having led the proposal and protocol development. SL contributed methodological expertise to the protocol’s development. EG, the primary supervisor, conceived the study, contributed to its design, and developed and drafted the study proposal and protocol. All authors were involved in further conceptualising and designing the study and have approved the submitted version of the manuscript. All authors agree to be personally accountable for their contributions and to ensure that any questions regarding the accuracy or integrity of any part of the work, even those they were not directly involved in, are appropriately investigated, resolved, and documented in the literature. Conceptualisation: GDA, SL, DG, KM, and EG. Methodology: GDA, SL, and EG. Resources: GDA, SL, DG, KM, and EG. Writing—original draft preparation: GDA, SL, and EG. Writing—review and editing: GDA, SL, DG, KM, and EG. Supervision: SL, DG, KM, and EG.

## Ethics

This project has received ethical approval from the University College London (UCL) Research Ethics Committee, identified by Project ID 27289/001. Additional approval was granted by the KATH-IRB - Komfo Anokye Teaching Hospital Institutional Review Board (KATH-IRB), referenced as KATH IRB/AP/087/24. All study participants will be provided with written informed consent to participate.

## Acknowledgements

We would like to express our sincere gratitude to Professor Daniel Ansong and the SMAART Consortium Team in Ghana for their invaluable assistance with the KATH-IRB ethics application process.

## Abbreviations

COMET: Core Outcome Measures in Effectiveness Trials
COREQ: Consolidated Criteria for Reporting Qualitative Research
COS: Core Outcome Set
COS-STAP: Core Outcome Set-Standardised Protocol Items
KATH-IRB: Komfo Anokye Teaching Hospital Institutional Review Board
PIS: Participant information leaflets
SMAART: Severe Malaria Africa - A Consortium for Research and Trials
UCL: University College London
UK MRC: United Kingdom Medical Research Council
WHO: World Health Organization

